# Validation of a rapid, saliva-based, and ultra-sensitive SARS-CoV-2 screening system for a pandemic-scale infection surveillance

**DOI:** 10.1101/2021.11.04.21265951

**Authors:** Robert E Dewhurst, Tatjana Heinrich, Paul Watt, Paul Ostergaard, Jose Maria Marimon, Mariana Moreira, Philip E Houldsworth, Jack D Rudrum, David Wood, Sulev Kõks

## Abstract

Without any realistic prospect of comprehensive global vaccine coverage and lasting immunity, control of pandemics such as COVID-19 will require implementation of large-scale, rapid identification and isolation of infectious individuals to limit further transmission. Here, we describe an automated, high-throughput integrated screening platform, incorporating saliva-based loop-mediated isothermal amplification (LAMP) technology, that is designed for population-scale sensitive detection of infectious carriers of SARS-CoV-2 RNA. Central to this surveillance system is the “Sentinel” testing instrument, which is capable of reporting results within 25 minutes of saliva sample collection with a throughput of up to 3,840 results per hour. It incorporates continuous flow loading of samples at random intervals to cost-effectively adjust for fluctuations in testing demand. Independent validation of our saliva-based RT-LAMP technology on an automated LAMP instrument coined the “Sentinel”, found 98.7% sensitivity, 97.6% specificity, and 98% efficiency against a RT-PCR comparator assay, confirming its suitability for surveillance screening. This Sentinel surveillance system offers a feasible and scalable approach to complement vaccination, to curb the spread of COVID-19 variants, and control future pandemics to save lives.

**One-Sentence Summary:** Development of a high-throughput LAMP-based automated continuous flow, random access SARS-CoV-2 screening platform with sufficient sensitivity and specificity to enable pandemic-scale population testing of infectious individuals using saliva sampling.

## Introduction

The SARS-CoV-2-induced COVID-19 pandemic has overwhelmed many healthcare systems, economies, and societies globally. Most countries are still struggling to control the spread of the disease more than two years since the emergence of the pandemic. By the 20th of February 2022, at least 423 million cases had been recorded cumulatively worldwide, with more than 5.8 million deaths (https://coronavirus.jhu.edu/map.html). Even by the middle of 2020, 90% of infections were still not captured by surveillance systems ^1^, leading to a gross underreporting of prevalence. Based on mortality rates, it was estimated that only 1-2% of COVID-19 cases were initially detected ^2^, while underlying SARS-CoV-2 infection being confirmed in only 30% of cases was attributed to excess mortality in the US ^3,4^. Clearly, epidemiological viral surveillance measures were not sufficiently effective in halting the rapid spread of infection globally.

While there is a wide range in the estimated presymptomatic and asymptomatic prevalence of SARS-CoV-2, many SARS-CoV-2-infected persons never go on to develop symptoms ^1,5,6^ or only do so after being infectious for several days. Even before emergence of the delta variant, an extensive systematic review and meta-analysis found that the average asymptomatic prevalence of SARS-CoV-2 infection was 17% (within a reported range of 4-52% ^7^). Since presymptomatic and asymptomatic individuals often vary considerably in the load of SARS-CoV-2 virus they carry, surveillance strategies should ideally have sufficient sensitivity to identify all potentially infectious individuals. This will be critical for identifying ‘superspreaders’, who have been estimated to account for up to 40% of subsequent infections, despite not necessarily having particularly high viral loads in their bodily fluids (including saliva). ‘Superspreaders’ are instead postulated to spread the virus more rapidly by alternative means, such as increased aerosol production ^8,9^.

The combined presymptomatic and asymptomatic infection rate has been estimated to exceed 50% in some unvaccinated populations, meaning that testing only symptomatic individuals will fail to detect many SARS-CoV-2 diagnoses. In the UK, 70% of cases were reported as undetected (8), while a French study found that 93% of infected persons were undetected in a single test ^1,5^. A comprehensive and detailed study from the Vo municipality in Italy, where 80% of the population was tested twice for the virus, found that 42% of those infected did not show any symptoms ^10^. A more recent report revealed that as much as 59% of all SARS-CoV-2 viral transmission came from asymptomatic transmission events (comprising 35% from presymptomatic individuals and 24% from individuals who never went on to develop COVID-19 symptoms) ^11^.

While vaccination greatly reduces the degree of asymptomatic virus carriage, a recent study showed that viral RNA from SARS-CoV-2 ‘delta’ variant breakthrough infection cases (79% of whom were asymptomatic) could still be detected in vaccinated individuals for up to 33 days (median 21 days) from their original diagnosis ^12^. This study found that delta viral loads were 251 times higher than viral loads reported from infections of previous SARS-CoV-2 strains detected almost a year earlier from the same region using equivalent RT-PCR tests ^12^. The increased infectiousness of the delta variant may ultimately be due to these much higher peak viral loads, which were shown to peak within 2-3 days on either side of the time of symptom development. These findings suggest that a more comprehensive vigilance strategy may be required to rapidly detect infectious individuals in the community, irrespective of whether they have symptoms, and regardless of vaccination status, to effectively contain the spread of the more infectious delta and omicron variants into vulnerable populations or populations with low prevalence.

The main bottlenecks with current testing strategy are limited capacity, route of the sampling and time that is required for testing. Current standard of testing involves RT-PCR analytics, that will take at least 2 hours to run from purified RNA. We need to add time that is required to take nasopharyngeal swab and the limit of skilled personnel who can safely do so. After taking sample, RNA needs to be extracted and purified, followed by the setting up and running the reaction. The best time from sampling to testing with RT-PCR would be 6 hours, in real-life this takes even longer. Capacity limitations are caused by the number of machines available in testing laboratories and by the limited number of personnel that can take and process the samples. Delays in the sampling, testing, and reporting the test results cause the escape of infectious individuals from surveillance. Therefore, the testing speed can be considered the biggest challenge with current pandemic.

To address sampling bottlenecks, we developed a saliva-based testing system that is robust, sensitive and easy to perform from multiple parallel self-sampling stations, The testing procedure begins with automated self-administered barcode ID scanning and saliva sampling via a small swab in less than 30 seconds per scanning point. Saliva sample tubes can then be continuously loaded onto a conveyor reflow oven for heated inactivation within 10 minutes, before parallel automatic tube decapping and transfer of samples into a reverse transcription loop-mediated isothermal amplification (RT-LAMP) assay incubation for 20-25 minutes.

We developed an automated random access, continuous flow robotic system known as the Sentinel for scaling the entire process of sample dispensing, incubation, and detection of RT-LAMP reactions while securely reporting results in up to 35 minutes including sample preparation. The scalable screening capability is intended to be rapidly deployed at low cost for regular surveillance testing of large numbers of individuals (particularly at borders, ports, and public sporting and entertainment venues). This surveillance system is also intended to enable rapid containment of emerging SARS-CoV-2 variants (omicron) that might eventually escape vaccine protection.

## Results

Here, we report a proof-of-concept pilot for the integration of all the required components of such a testing system, incorporating saliva sample preparation, optimized reverse transcriptase-loop mediated isothermal amplification (RT-LAMP) chemistry and the development of a random access, continuous flow system for scaling the entire process of sample dispensing, incubation, and detection of reactions while securely reporting results in real time.

We chose to employ RT-LAMP chemistry for rapid virus detection, since its sensitivity is much closer to RT-PCR than to lateral flow assay (LFA)/ lateral flow device (LFD) or rapid antigen tests (RAT) ^13,14^ and because RT-LAMP enzymes are relatively tolerant of inhibitors present in saliva, allowing readout of accurate test results within minutes of direct sample collection ^15^.

### Identifying robust and sensitive RT-LAMP chemistries for surveillance screening

We first performed a side-by-side comparison of RT-PCR against a range of fluorometric RT-LAMP chemistries utilising five published primer sets and using RNA extracted from 20 clinical saliva samples obtained from individuals who had received a positive nasopharyngeal swab RT-PCR test result, including 10 samples with Ct above 34 indicating very low viral loads. For this evaluation, we selected the most sensitive RT-PCR assay in the FDA’s published list of 117 SARS-CoV-2 tests (limit of detection (LoD): 180 viral copies/ml) as our benchmark comparator assay ^16^.

When the same amount of RNA was added to each assay to allow a fair and direct comparison, RT-LAMP detected SARS-CoV-2 virus in 10 out of 20 samples versus 11 out of 20 for RT-PCR all these samples with Ct at least 33 or less. This result indicates that RT-LAMP has comparable performance to RT-PCR with the same amount of RNA input (**Table 1**). We also found good concordance between the time-to-threshold (TTT) values across these RT-LAMP chemistries and cycle threshold (Ct) values in RT-PCR (**Suppl. Figure S1**). The Zhang E1/N2 primer set ^17^ was the highest performer out of the five primer sets tested in this comparison. This finding led us to include an alternative E1/N2 primer set, which is available commercially for clinical diagnostic use in Europe (Hayat Genetics), in subsequent comparative assays.

**Table 1.** Comparison of RT-LAMP and RT-PCR results from extracted RNA from clinical saliva samples. 2 μl of purified RNA from 20 clinical saliva samples were used for each assay type to provide a fair comparison. RT-LAMP primer sets are ordered (left-to-right) from highest to lowest performer. Performance was measured by the highest Ct below which 100% sensitivity was achieved. Samples are ordered (top-to-bottom) by viral load, from lowest Ct to highest Ct. PE kit: PerkinElmer New Coronavirus Nucleic Acid Detection Kit. TTT: time to threshold, where threshold is defined as 1.5x baseline. Dash (-) denotes either no detection of target by 45 cycles of RT-PCR or fluorescence signal did not reach threshold by 30 min in LAMP.

|  |  | RT-qPCR on extracted RNA |  |  | RT-LAMP on extracted RNA |  |  |  |  |  |
| --- | --- | --- | --- | --- | --- | --- | --- | --- | --- | --- |
| Chemistry |  | Ct values; RT-qPCR (PE kit); 2 µl RNA |  |  | TTT (min); RT-LAMP (NEB E1700 + Syto9); 2 µl RNA |  |  |  |  |  |
| LAMP primer set(s) |  |  |  |  | Zhang E1/N2 | Huang O117 | Zhang E1 | Novacyt-S | Novacyt-S | Huang S17 |
| Target |  | N | ORF1ab | Lowest | N/E | ORF1ab | E | S | S | S |
| 10 |  | 24.7 | 23.9 | 23.9 | 14.0 | 8.0 | 10.5 | 13.5 | 14.5 | 9.5 |
| 1 |  | 28.9 | 28.6 | 28.6 | 17.5 | 9.5 | 11.5 | 16.5 | 16.5 | 10.0 |
| 14 |  | 31.3 | 31.2 | 31.2 | 18.0 | 11.0 | 14.0 | 19.5 | 19.0 | 11.5 |
| 7 |  | 32.6 | 32.8 | 32.6 | 19.5 | 10.0 | 14.0 | 19.5 | 22.5 | - |
| 6 |  | 33.0 | 34.1 | 33.0 | 21.0 | 11.5 | 14.5 | 25.0 | - | 13.0 |
| 5 |  | 34.6 | 36.1 | 34.6 | 20.5 | 14.0 | 15.5 | 20.0 | - | 14.5 |
| 18 |  | 35.0 | 35.7 | 35.0 | 23.5 | 12.5 | 15.0 | - | - | 14.0 |
| 17 |  | 35.3 | 35.6 | 35.3 | 23.0 | 11.5 | - | - | - | 17.5 |
| 12 |  | 35.7 | 36.2 | 35.7 | 22.0 | - | 22.0 | - | - | 15.0 |
| 19 |  | 36.4 | 43.8 | 36.4 | - | - | 14.0 | 20.5 | - | - |
| 8 |  | - | 43.7 | 43.7 | - | - | 18.0 | - | - | - |
| 2 |  | - | - | - | - | - | - | - | - | - |
| 3 |  | - | - | - | - | - | - | - | - | - |
| 4 |  | - | - | - | - | - | - | - | - | - |
| 9 |  | - | - | - | - | - | - | - | - | - |
| 11 |  | - | - | - | 22.5 | - | - | - | - | - |
| 13 |  | - | - | - | - | - | 20.0 | - | - | - |
| 15 |  | - | - | - | - | - | - | - | - | - |
| 16 |  | - | - | - | - | - | - | - | 22.0 | - |
| 20 |  | - | - | - | - | - | - | - | - | - |

### Optimising saliva sample preparation

Large-scale surveillance screening applications requiring faster turn-around may not be compatible with the time or equipment required for the RNA purification. A variety of rapid protocols have been developed to address this challenge, such as ‘Saliva Direct’, which can detect 6-12 copies of SARS-CoV-2 per μl of saliva ^18^. However, in practice, PCR-based assays have generally proven to be too demanding for the rapid isolation of RNA from potentially infectious people visiting crowded high-risk exposure sites such as airplanes. While the literature increasingly supports the use of saliva samples for large-scale surveillance screening ^19,20^, we found saliva testing less straightforward in practice, requiring optimization of collection buffer and heat treatment to ensure robust and sensitive detection using RT-LAMP assays.

We therefore optimised sample preparation to allow rapid, sensitive, and repeatable testing directly from saliva without purification of RNA (direct RT-LAMP). To avoid time-consuming RNA extractions, we trialled a range of different heat inactivation protocols ^21,22^ and saliva collection solutions ^23,24^, all of which have been found to improve sensitivity in RT-PCR or RT-LAMP assays. However, we adopted a protocol in which fresh or frozen saliva was diluted at least 1 in 4 in AviSal Sample Collection Buffer (Hayat Genetics), followed by virus heat inactivation at 95°C for 10 minutes. This protocol was found to maintain the sensitivity of virus detection and allowed for stable storage of the samples at room temperature (see stability data reported below). To make this protocol compatible with our high-throughput surveillance system, we refined it by collecting saliva into 96-format tubes prefilled with AviSal. Sample tubes then transit a fan forced reflow oven for the heat inactivation step. These preprocessed samples are then ready to be analysed within minutes following tube racking and automated uncapping.

### Assessing the performance of direct RT-LAMP for surveillance screening

Using this optimized sample inactivation method, we next compared various dual read-out (fluorometric and colorimetric) direct RT-LAMP chemistries with two approved RT-PCR diagnostic tests, each requiring RNA to be extracted, and with two direct RT-PCR tests, on the same 20 clinical saliva samples (**Table 2**). Direct RT-LAMP detected virus in 15 out of 20 samples, compared to 17 and 18 out of 20 for the two RT-PCR diagnostics assays (**Table 2A**). The best RT-LAMP assays could detect viruses in all samples corresponding to a Ct of 33/34 and below, where Ct values were derived from an approved diagnostic comparator test (**Table 2B**). Encouragingly, detection of samples with low viral loads (Ct >40) was also observed, albeit with less than 100% sensitivity. The performance of direct RT-LAMP was comparable to that of direct RT-PCR, with direct RT-LAMP detecting virus in 15 out of 20 samples, versus 14 or 16 (depending on the PCR kit used) out of a total of 20 with direct RT-PCR (**Table 2A**). The detection limit to achieve 100% sensitivity of direct RT-LAMP versus direct RT-PCR corresponded to a Ct of 40 and below (**Table 2B**). Both primer sets targeting the N and E genes (in particular, the CE-marked Hayat Genetics diagnostic kit) performed better than the ORF1ab-targeted Huang O117 primer set ^25^. Time-to-threshold (TTT) values for all RT-LAMP assays performed were between 8.5 and 26.5 minutes, with a mean TTT of 15.1 min. Taken together, these data show that our direct RT-LAMP method (using optimized saliva inactivation and Hayat Genetics chemistry) is rapid and sufficiently sensitive to detect infectious levels of SARS-CoV-2 virus directly from as little as 1.25 μl of saliva diluted in AviSal buffer.

**Table 2.**
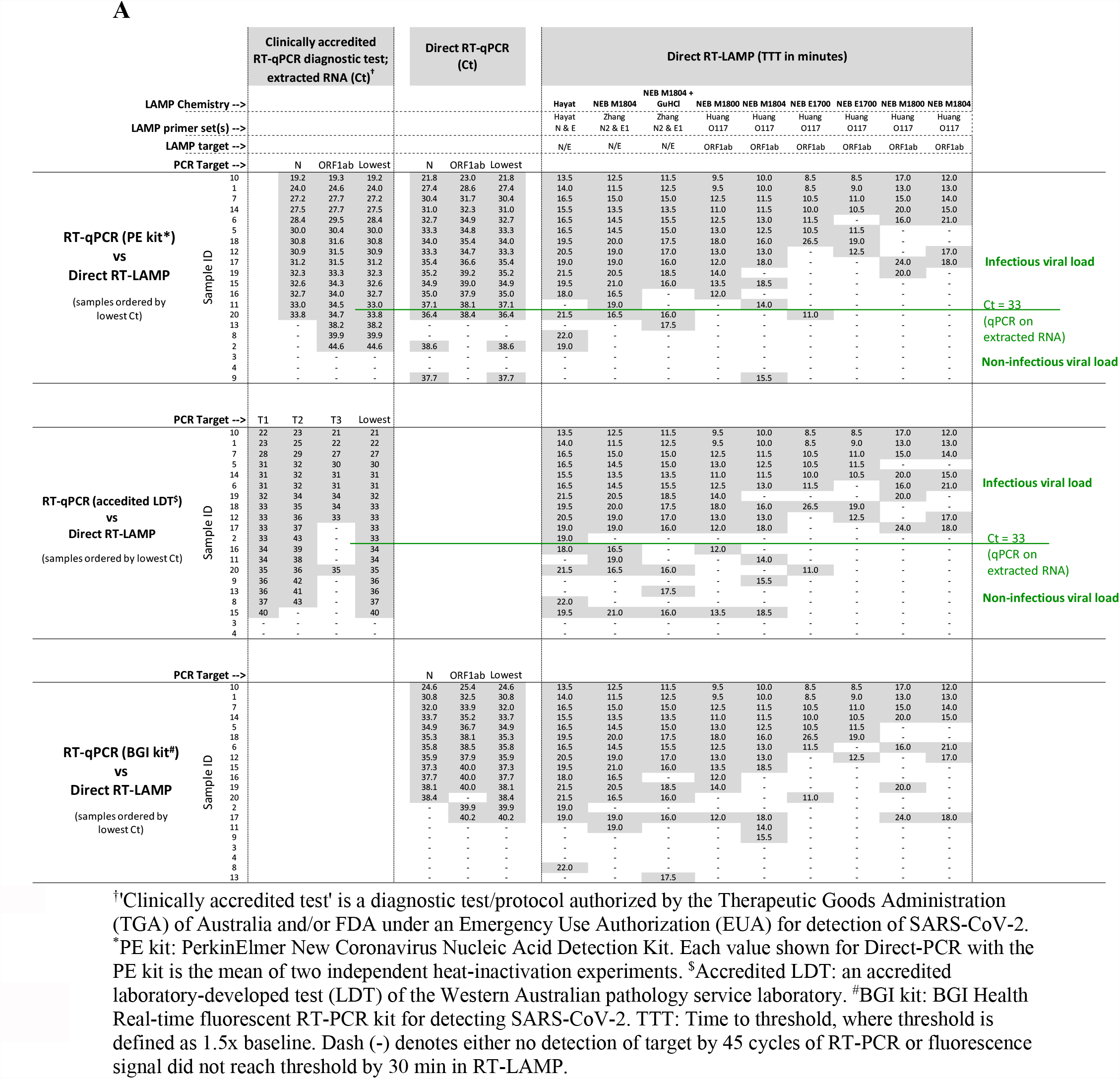
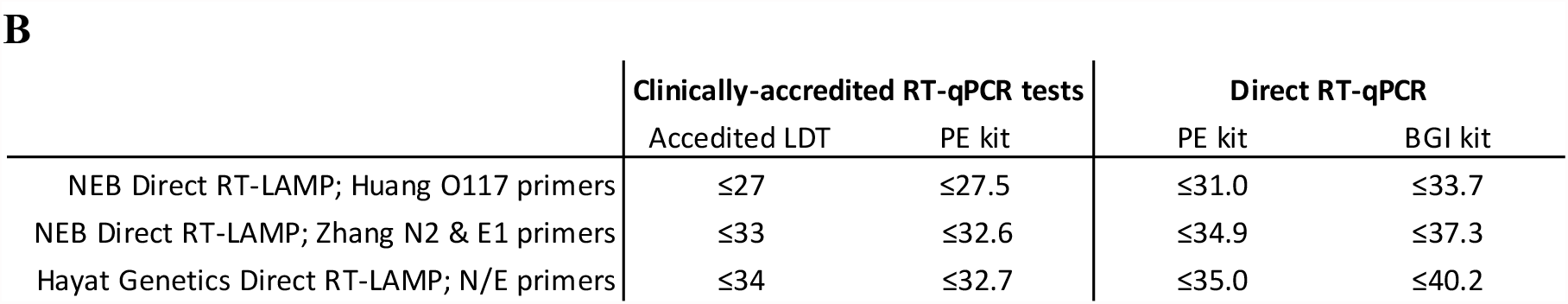
Performance of Direct RT-LAMP. A. Performance of Direct RT-LAMP compared to RT-PCR. B. Summary of RT-LAMP performance compared to RT-PCR tests: detection limits for 100% sensitivity. In Table A, RT-LAMP experiments are ordered (left-to-right) from highest to lowest performer. Performance was measured by the highest Ct below which 100% sensitivity was achieved. All NEB chemistries included added Syto9 for detection by fluorometry. All chemistries, except NEB E1700, were colorimetry capable. Colour changes were consistent with fluorometry results. The percentage of reaction volume consisting of sample in RT-LAMP and direct RT-PCR varied from 3.75% to 5%, according to the specified requirements of each test. Purified RNA input of the Perkin Elmer RT-PCR assay kits constituted two-thirds of the total reaction volume, according to the instructions for use.

Inconsistent detection of samples with very low viral loads would not be expected to be relevant to COVID-19 containment measures, since those samples likely correspond to individuals thought not to be contagious ^26^. If we consider only those positive samples with Ct values below 33 (green line in **Table 2A**) from potentially infectious individuals, then there is 100% concordance between RT-PCR tests and RT-LAMP targeting the N and E genes.

Our assay validation studies demonstrated that a variety of RT-LAMP chemistries can be used to detect the SARS-CoV-2 virus directly from as little as 1.25 μl of saliva input diluted in AviSal buffer with high sensitivity across samples spanning a wide range of viral loads (Ct values ranging from 19 to >40) when compared with RT-PCR “gold standard” comparator assays for the detection of SARS-CoV-2 (**Table 2**). Importantly, using the optimized RT-LAMP assay system, we observed 100% sensitivity in detecting SARS-CoV-2 across all specimens with potentially contagious viral loads, corresponding to Ct values up to 33. These data suggest that our surveillance system is likely to be sufficiently sensitive to detect all infectious SARS-CoV-2 carriers, given that an individual’s viral load beneath our limit of detection is generally thought not to be contagious, given multiple reported failures to cultivate any SARS-CoV-2 virus using *in vitro* cell culture from samples with Ct values of over minimum reported thresholds ranging from 24 to 33 ^26-28^. Indeed, sequencing of SARS-CoV-2 transcripts from infected cells recently established that full genome sequence coverage was not observed in samples with a Ct greater than 32 (measured with the same Perkin Elmer RT-PCR assay which we used here as our comparator ^29^), suggesting that any RT-PCR detection of small SARS-CoV-2 genomic fragments in samples beyond this threshold would not likely reflect intact viable virus with potential to be infectious.

### Optimising RT-LAMP specificity to minimise false positive reactions

Sequencing of products of LAMP reactions has demonstrated that this amplification chemistry can be prone to the development of false positives in the absence of template due to nonspecific primer-dependent amplification. These nonspecific reaction products, the prevalence of which varies between primer sets, normally emerge later in the reaction incubation ^30^, explaining why the ‘time-to-positive’ relates to specificity as well as to viral load.

Maximum incubation times are therefore typically specified in RT-LAMP protocols to minimise the chance of such false positives arising, even at the expense of reduced sensitivity. We found that in the absence of a sealed heated lid on the Sentinel surveillance instrument, a high rate of evaporation during a standard 30-minute RT-LAMP incubation at 65°C resulted in the early emergence of false positives after approximately 20 minutes, causing the reaction specificity to drop below 100%. This finding is particularly relevant to screening on the Sentinel surveillance instrument (see below), which lacks a heated lid seal to minimise evaporation. To better understand the correlation between the rate of false positive production and evaporation, we ran multiple negative control colorimetric RT-LAMP reactions in replicate, with both NEB (M1804 plus E1/N2 primers) and Hayat Genetics chemistries using saliva samples negative for SARS-CoV-2 and extended the reaction run time beyond the standard 30 min to 90 minutes (**Figure 1**). In the absence of mineral oil, 10% of NEB reactions were positive by 30 min, with 100% becoming positive by 40 minutes. The more specific Hayat Genetics assay chemistry was slower to produce false positives, with all reactions still negative at 40 minutes but eventually turning positive by 70 min in the absence of mineral oil. These data confirm that RT-LAMP is prone to false positives and that choice of optimal chemistry and preventing evaporation is critical in ensuring high specificity of RT-LAMP by 30 minutes. The Hayat Genetics assay also exhibited superior specificity. However, to further mitigate the effect of evaporation, regardless of the chemistry used, we incorporated an oil overlay barrier in all RT-LAMP reactions.

**Fig. 1.**
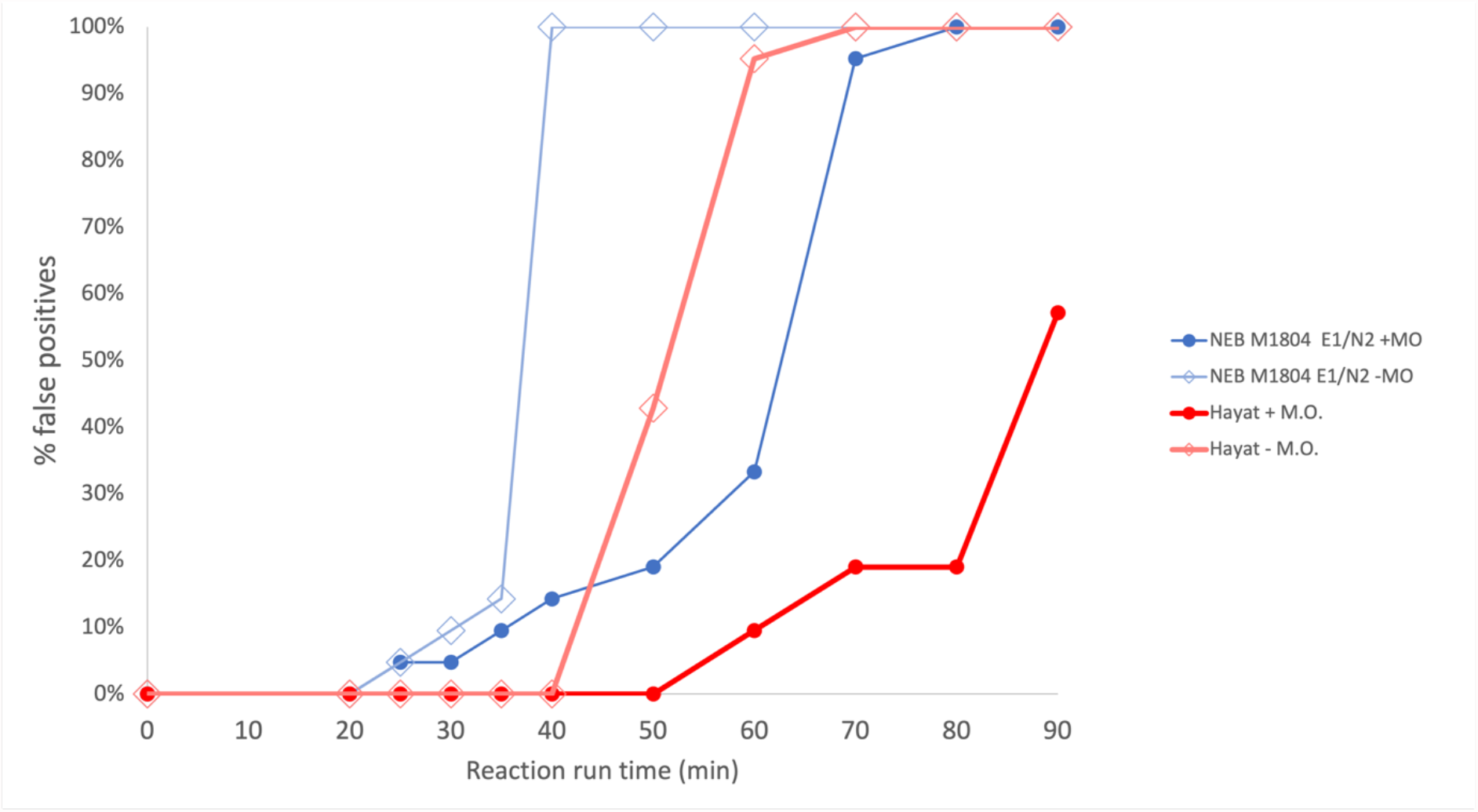
The presence of mineral oil markedly reduces the rate of production of false positive RT-LAMP reactions. Two assay chemistries were compared: NEB WarmStart Colorimetric LAMP with UDG (M1804) and Hayat Rapid Colorimetric & Fluorometric One Step LAMP SARS-CoV-2 Test Kit, each set up with and without 15 μl mineral oil overlay. Twenty-one identical replicate negative control reactions were set up per condition with a single saliva sample negative for SARS-CoV-2 diluted in VTM and AviSal at a ratio of 1:1:2 and heat inactivated for 10 min at 95°C. The sample was added to give 5% (NEB) and 3.75% (Hayat) final concentrations of crude saliva in a 25 μl reaction volume. + M.O. with mineral oil overlay; - M.O. without mineral oil. In a typical 30-minute reaction runtime, only Hayat chemistry resulted in no false positives and 100% specificity. For the NEB chemistry, false positives were observed after 20 minutes, even with a mineral oil overlay. Mineral oil overlay markedly reduces the false positive rate.

### Confirmation of the detection of “Delta” and “Omicron” variants by RT-LAMP

We next investigated whether the most effective of the RT-LAMP chemistries we tested (Hayat Genetics) could detect RNA from the SARS-CoV-2 “delta” (B.1.617) and “omicron” (B.1.1.529) variants. As expected, given that the Hayat RT-LAMP primer set amplifies genomic regions that are conserved across variants, we could robustly detect synthetic RNA from the delta and omicron variants across a wide range of concentrations within 20 minutes (**Suppl. Figure S2A and S2B**).

### Designing an RT-LAMP surveillance system supporting continuous loading of samples at random

We designed an automated RT-LAMP surveillance system, specifically for ultra-high-throughput detection of viral nucleic acids, directly from saliva samples taken on a population-wide scale (**Figure 2**). This integrated system combines an optimized system for efficient sample collection preparation in barcoded tubes, which are rapidly heat inactivated and consolidated into racks for automated uncapping and continuous loading onto a robotic instrument that automatically dispenses samples and reagents and continuously scans and reports results.

**Fig. 2.**
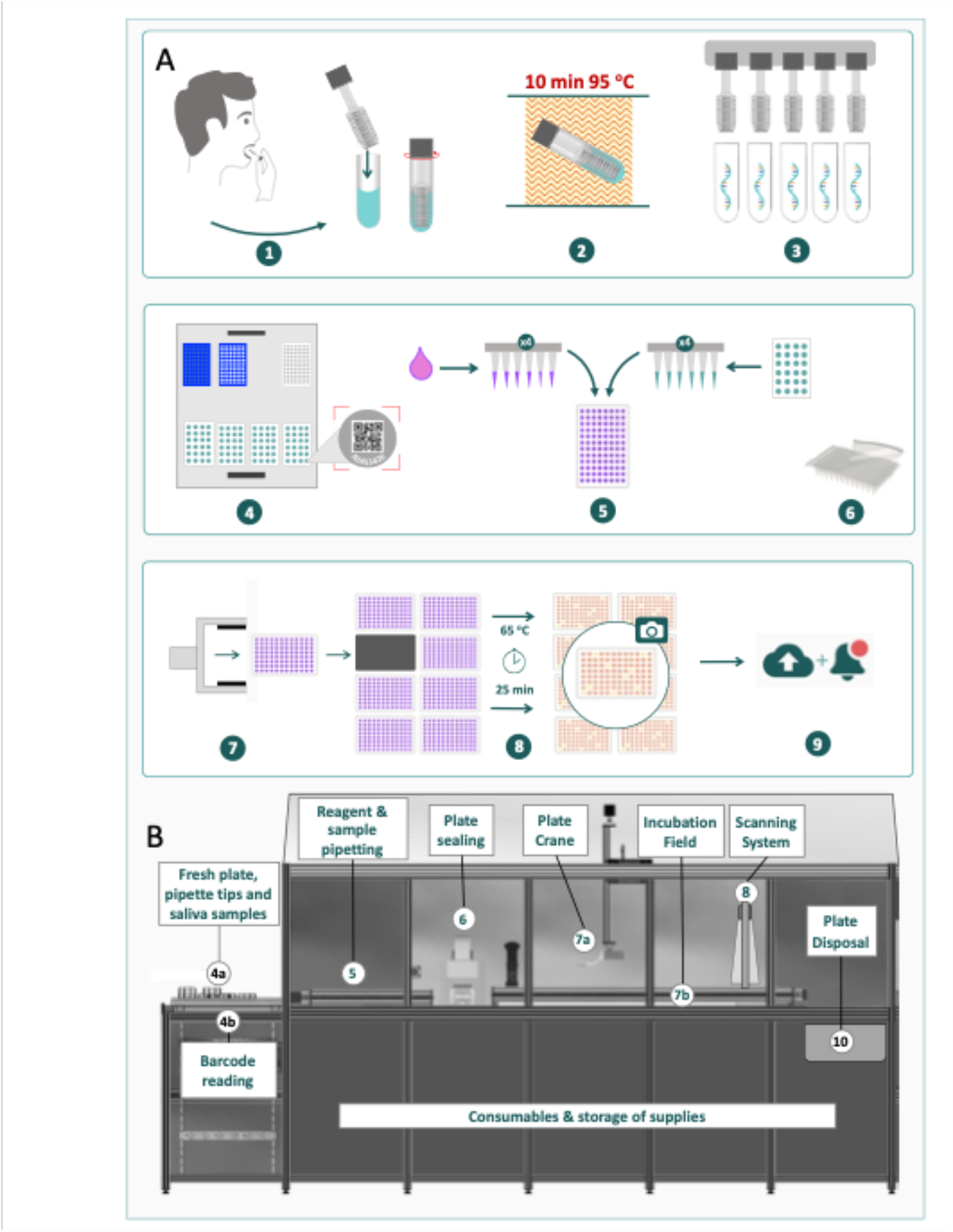
Process flow from saliva collection to data reporting. A: Schematic diagram of the main steps of the process flow. B: Sentinel robot for LAMP reaction setup, incubation, and detection. Individual process steps: 1. Saliva collection via microswabs into test tubes containing inactivation solution. 2. Sample tubes with saliva dilutions were heat-inactivated for 10 minutes at 95°C. 3. Automated uncapping of a complete sample tube rack. 4. Sample tube barcode reading during Sentinel robot loading. 5. The automated pipetting system adds a selected sample volume to the RT-LAMP master mix in clear reagent plates. 6. Plate sealing by automated heat sealer. 7. Plate crane transport of the reaction plate onto a 65°C incubation field. 8. LAMP reactions were incubated at 65°C for 25 min; digital, parallel scanning of multiple 96-well microplates. 9. Real-time, secure reporting of deidentified data and analysis. 10. Disposal of scanned microplates.

The integrated system was designed to address key bottlenecks identified in existing surveillance technologies by incorporating the following features:

i. A scalable method for safe collection, heat inactivation and tracking of saliva samples.
ii. Optimized sample processing and LAMP assay choice to maximise sensitivity/specificity.
iii. Incorporation of a continuous flow, random access loading of racks of sample tubes/plates onto the system with integrated sample tracking and reporting to support ultra-high throughput applications.

The capacity of conventional molecular diagnostic instruments is constrained by largely batch-based sample loading logistics, frequently resulting in rate-limiting queuing of microplates awaiting access to the instrument. This batch constraint also applies to large ‘continuous flow’ diagnostic instruments ^31^, which sometimes include multiple incubation stations employed on a range of predetermined schedules to achieve faster cycle times, which can be less time- and cost-efficient, particularly at lower throughputs, while samples are accumulated to achieve optimal capacity.

To address this scalability bottleneck, the ‘Sentinel’ LAMP instrument (**Figure 2B)** was designed as a truly continuous flow system, treating the arrival and disposal of each plate into the system completely independently. The Sentinel instrument can perform RT-LAMP tests at up to 3,480 tests per hour, with potential for further increases in scale in the future, by switching from a 96- to a 384-well format.

The Sentinel system was conceived to incorporate a resource scheduling and monitoring system tasked with processing as many microplate assays as possible within a given period, balancing upstream processing of samples with downstream reaction and disposal steps, to efficiently schedule the arrival and departure of microplates being analysed into selected incubation slots within a common isothermal incubation zone.

### Sample stability experiment: Hayat RT-LAMP assays (N & E targets)

To establish the feasibility of saliva sample collection without requiring cold chain logistics, we tested how our sample collection procedure could affect the outcome of RT-LAMP assays and challenged the robustness of testing with different storage conditions using AviSal saliva collection buffer. This study established that collected saliva samples spanning a wide range of viral loads are stable in storage media without loss of sensitivity during preinactivation storage at RT for up to 48 h, post inactivation at RT for 2 h or 4 °C for 48 h, and through a freeze/thaw cycle. (**Figure 3**).

**Fig. 3.**
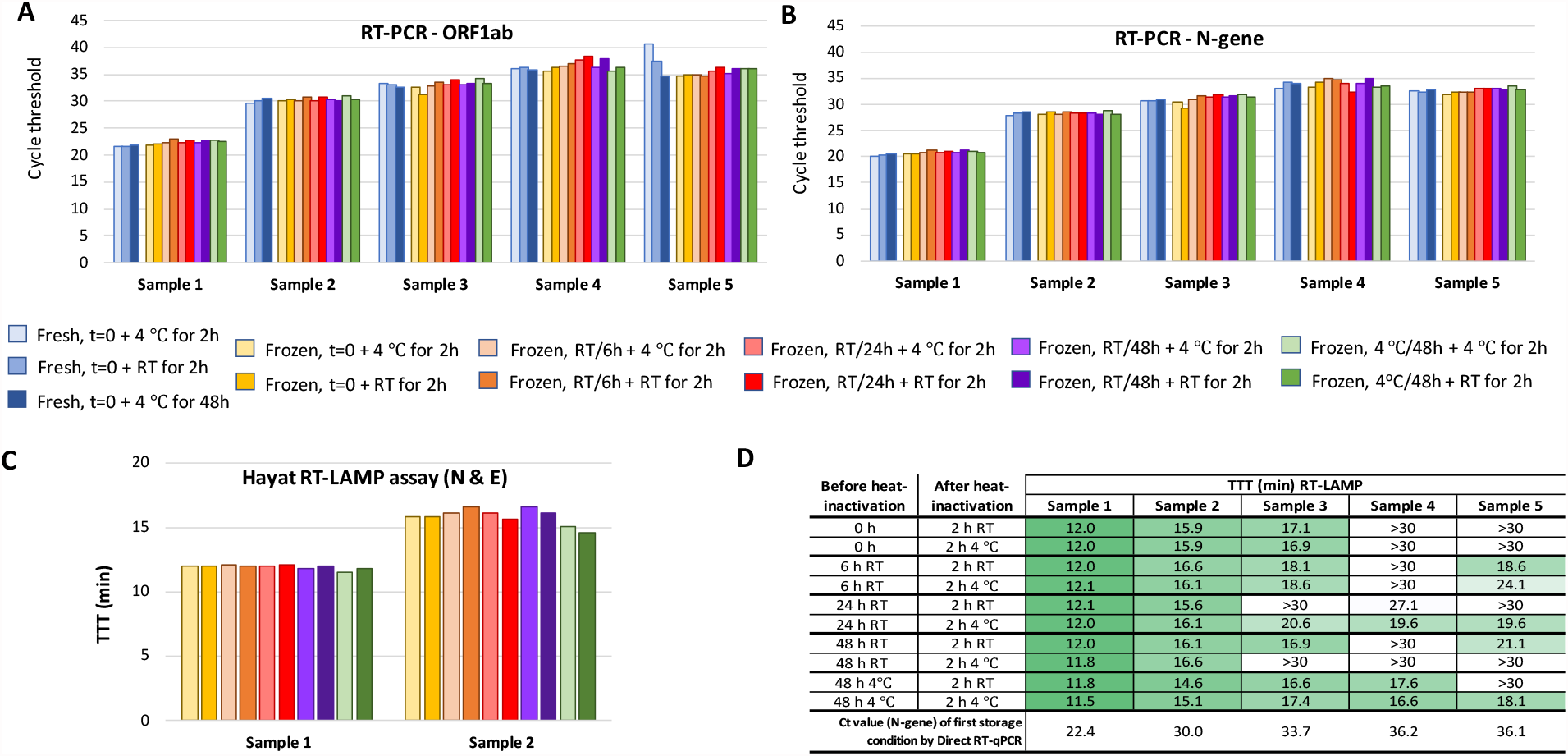
Stability of saliva samples during short-term storage determined by RT-PCR and RT-LAMP. Five sample pools were prepared from ten saliva samples to give a range of viral loads corresponding to predicted Ct values (from extracted RNA) from 24.6 to 35.0 for the N-gene and 25.8 to 39.1 for ORF1ab. Sample pools were diluted with an equal volume of AviSal buffer and stored short-term under the following test conditions prior to heat inactivation: room temperature (RT) for 0, 6, 24 and 48 hours (t=0, 6 h, 24 h & 48 h) or 4 °C for 48 hours. Samples were then heat inactivated at 95 °C for 10 min and frozen. Upon thawing, samples were stored post inactivation at either 4 °C or RT for 2 h or 4 °C for 48 h (t=0 only). The 4 °C incubation was taken as baseline as a proxy for no post-inactivation storage. RT-PCR (BGI Real-Time Fluorescent RT-PCR Kit for Detecting SARS-CoV-2), targeting ORF1ab (A) and the N gene (B), and RT-LAMP (Hayat Genetics) (C and D) assays were performed on samples subjected to each of these conditions. To understand the effect of freezing samples prior to heat inactivation, for the t=0 timepoint, a fresh versus frozen comparison was performed. Viral loads (as measured by RT-PCR on extracted RNA) of sample pools #3, 4 and 5 are too low for 100% sensitivity of detection by RT-LAMP – all have Ct ≥33 for N gene. Hence, not all reactions were positive within the 30 min runtime (D). All Ct and TTT values were calculated as the means of technical duplicates.

### Colorimetric LAMP limit of detection on the clinically deployed Sentinel instrument

Next, we conducted a study to pilot the implementation of the Sentinel Surveillance instrument in the context of a clinical microbiology service laboratory in San Sebastian, Spain. Saliva samples from individuals who had tested positive by an approved diagnostic RT-PCR swab test were frozen prior to testing on the Sentinel Instrument. Six of these saliva samples, with distinct viral loads, were serially diluted to obtain a panel of 36 samples spanning a wide range of Ct values. These diluted samples were further diluted in a commercially available saliva transport medium – Saliva Transport buffer M (Vitro SA, Spain) – and heat inactivated for 10 min at 95°C. Direct RT-PCR (Seegene Allplex) was performed on 5 μl of these heat-treated saliva dilutions.

The NEB colorimetric RT-LAMP (N&E-gene, M1800 2x LAMP mix) was run on the Sentinel using 3 μl of the same template as RT-PCR. **Table 3A** shows the results of RT-PCR and RT-LAMP for all 36 saliva samples organized by sample, while **Table 3B** shows the same data organized by Ct value. Table B clearly demonstrates that all saliva samples up to a Ct of 31.1 were detectable by RT-LAMP. Higher Ct values could also be detected, but with inconsistent reliability, indicating that the limit of detection for VTM/heat-treated saliva samples combined with NEB M1800/N&E chemistry corresponds to a Ct of 31.1, which represents a slightly less sensitive detection threshold than we obtained for similar chemistry with an alternative AviSal Sample Transport Buffer (**Table 2**). This decreased sensitivity observed may be due to differences in the composition of the alternative sample collection buffer used or, alternatively, could result from differences in sensitivity between colorimetric/fluorometric chemistries or differences in the comparator RT-PCR assays used in each case. Nevertheless, this pilot implementation study demonstrated that the SARS-CoV-2 RT-LAMP assay is sufficiently versatile to be adapted to be compatible with local sample collection processes of a routine clinical diagnostics service.

**Table 3.** RT-LAMP limit of detection in saliva samples carried out on a Sentinel instrument in a clinical pathology laboratory. Six SARS-CoV-2 saliva samples with different viral loads were serially diluted 1:10 in nuclease-free water down to 1:100,000 dilutions to obtain 36 samples with a wide range of Ct values. The diluted samples were further diluted 1:3 in Vitro Diagnostica Saliva Transport buffer M (VTM) and heat inactivated for 10 min at 95°C. Direct RT-PCR (Seegene Allplex) was carried out with 5 μl of heat-treated saliva dilution in VTM; NEB colorimetric RT-LAMP (N&E-gene, M1800 2x LAMP mix) was carried out on a Sentinel station with 3 μl of the same template as RT-PCR. Table A shows the results of RT-PCR and RT-LAMP for all 36 saliva samples organized by sample. Table B shows the same data organized by Ct value. Table B clearly demonstrates that all saliva samples up to a Ct of 31.1 were detected by RT-LAMP. Samples with higher Ct values started showing stochastic signals, indicating that the limit of detection for VTM/heat-treated saliva samples combined with NEB M1800/N&E chemistry is above a Ct of 31.

**A**
| Sample | RT-PCR [Ct] | RT-LAMP TTT [min] |
| --- | --- | --- |
| Pos 1 neat | 21.49 | 16 |
| Pos 1 E-01 | 23.26 | 17 |
| Pos 1 E-02 | 28.06 | 21 |
| Pos 1 E-03 | 32.79 | 22 |
| Pos 1 E-04 | 34.04 | 22 |
| Pos 1 E-05 | 38.17 | - |
| Pos 2 neat | 26.44 | 20 |
| Pos 2 E-01 | 28.22 | 20 |
| Pos 2 E-02 | 32.11 | 20 |
| Pos 2 E-03 | 37.72 | 21 |
| Pos 2 E-04 | nd | 23 |
| Pos 2 E-05 | 38.08 | - |
| Pos 3 neat | 23.29 | 19 |
| Pos 3 E-01 | 23.42 | 19 |
| Pos 3 E-02 | 29.79 | 22 |
| Pos 3 E-03 | 32.39 | - |
| Pos 3 E-04 | 35.67 | - |
| Pos 3 E-05 | nd | - |
| Pos 4 neat | 23.06 | 21 |
| Pos 4 E-01 | 27.15 | 22 |
| Pos 4 E-02 | 27.56 | 22 |
| Pos 4 E-03 | 32.79 | - |
| Pos 4 E-04 | 36.61 | - |
| Pos 4 E-05 | 37.54 | - |
| Pos 5 neat | 25.49 | 17 |
| Pos 5 E-01 | 25.28 | 17 |
| Pos 5 E-02 | 31.04 | 19 |
| Pos 5 E-03 | 34.26 | 20 |
| Pos 5 E-04 | 36.21 | - |
| Pos 5 E-05 | nd | - |
| Pos 6 neat | 28.33 | 23 |
| Pos 6 E-01 | 29.99 | 20 |
| Pos 6 E-02 | 32.79 | 23 |
| Pos 6 E-03 | nd | - |
| Pos 6 E-04 | nd | - |
| Pos 6 E-05 | nd | - |

**B**
| RT-PCR [Ct] | RT-LAMP TTT [min] |
| --- | --- |
| 21.49 | 16 |
| 23.06 | 21 |
| 23.26 | 17 |
| 23.29 | 19 |
| 23.42 | 19 |
| 25.28 | 17 |
| 25.49 | 17 |
| 26.44 | 20 |
| 27.15 | 22 |
| 27.56 | 22 |
| 28.06 | 21 |
| 28.22 | 20 |
| 28.33 | 23 |
| 29.79 | 22 |
| 29.99 | 20 |
| 31.04 | 19 |
| 32.11 | 20 |
| 32.39 | - |
| 32.79 | 22 |
| 32.79 | - |
| 32.79 | 23 |
| 34.04 | 22 |
| 34.26 | 20 |
| 35.67 | - |
| 36.21 | - |
| 36.61 | - |
| 37.54 | - |
| 37.72 | 21 |
| 38.08 | - |
| 38.17 | - |
| nd | 23 |
| nd | - |
| nd | - |
| nd | - |
| nd | - |
| nd | - |

### Independent validation, specificity, and sensitivity of the saliva-based LAMP assay

The independent laboratory performed validation of Avicena RT-LAMP testing system by comparing RT-PCR results of 150 COVID-19 positive and 250 negative samples. This comparative analysis indicated that Avicena RT-LAMP detection has a sensitivity of 98.7% (95.3-99.6%) and a specificity of 97.6% (94.8-98.9%) against a RT-PCR comparator standard with the cut-off time to TTT was set to 22 min (Table 4). The efficiency of the test was 98%. Changing the maximum TTT cut-off time altered the sensitivity and the specificity of the test (Table 4). Shorter TTT cut-off (20 minutes) resulted in better specificity (99.6%) and in lower sensitivity (95.3%). TTT cut-off values were in a perfect correlation with the Ct values of the RT-PCR results showing (Supplementary Figures S3 and S4).

**Table 4.** Hayat Genetics RT-LAMP sensitivity, specificity, positive predictive value (PPV), negative predictive value (NPV) and efficiency (A). Four different TTT cut-off times were used for comparison with RT-PCR results. Sensitivity and specificity with 95% confidence limits (B).

**A**
| Parameter | Cut-off (mins) |  |  |  |
| --- | --- | --- | --- | --- |
|  | 20 | 21 | 22 | 23 |
| Sensitivity | 95.3 | 96.0 | 98.7 | 99.3 |
| Specificity | 99.6 | 98.4 | 97.6 | 95.2 |
| PPV | 99.3 | 97.3 | 96.1 | 92.5 |
| NPV | 97.3 | 97.6 | 99.2 | 99.6 |
| Efficiency | 98.0 | 97.5 | 98.0 | 96.7 |

**B**
| Parameter | Cut-off (mins) |  |  |  |
| --- | --- | --- | --- | --- |
|  | 20 | 21 | 22 | 23 |
| Sensitivity | 95.3 | 96.0 | 98.7 | 99.3 |
| Lower 95% | 90.7 | 91.5 | 95.3 | 96.3 |
| Upper 95% | 97.7 | 98.2 | 99.6 | 99.9 |
| Specificity | 99.6 | 98.4 | 97.6 | 95.2 |
| lower 95% | 97.8 | 95.9 | 94.8 | 91.8 |
| upper 95% | 99.9 | 99.4 | 98.9 | 97.2 |

We also compared the performance of Avicena RT-LAMP test to other approved testing solutions, direct RT-LAMP and RAT/LFA/LFD test. The Hayat RT-LAMP assay run on an Avicena Sentinel instrument (designated as Hayat/Avicena test) performed better that direct RT-LAMP (Optigene) run on the recommended Genie instrument or on the Sentinel instrument (Figure 4). The Hayat/Avicena test worked much better that LFA/LFD/RAT test run with the recommended saliva protocol (Figure 4). The RAT test showed very poor performance with only 50% of detection of high viral load samples.

**Fig. 4.**
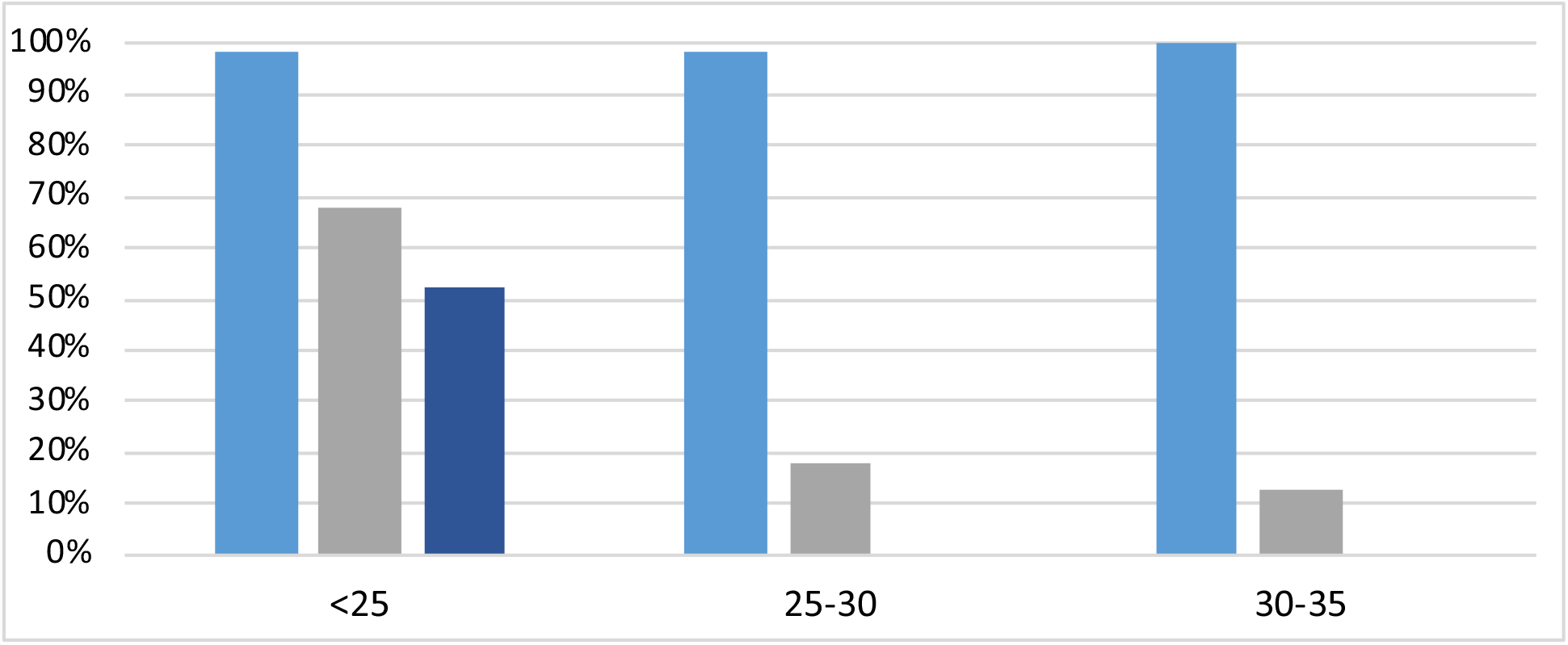
Comparison of sensitivity of different assay technologies grouped by Ct values. Five Light blue - Hayat/Avicena test, grey – direct RT-LAMP, dark blue – RAT/LFD/LFA. Direct LAMP test had initial sensitivity 68% and lost it significantly from Ct 25. RAT/LFD/LFA tests had initial sensitivity 50% and lost it completely from Ct 25 and above.

## Discussion

### Addressing the need for more sensitive rapid surveillance of asymptomatic individuals

While the diagnostic gold standard RT-PCR test is generally sufficiently sensitive to detect all infected individuals, this technology lacks sufficient speed (taking hours) and throughput (at most a few thousand samples processed per instrument per day) to support routine surveillance screening and quarantine applications. In addition, nasopharyngeal sampling requires trained personnel, is inconvenient, and has decreased participant acceptance if multiple testings within a short period are required.

Alternatively, more acceptable rapid tests are emerging in response to these challenges but are not all sufficiently sensitive to reliably prevent outbreaks, particularly as more infectious variants continue to arise. A recent systematic review suggested an average analytical sensitivity overall for lateral flow antigen (LFA) tests of 72% ^32-34^, while a systematic review of 58 LFA test evaluations found that their sensitivity was reliable only when detecting samples with high (i.e. Ct<25) viral loads. This explains why their sensitivity was significantly lower in asymptomatic people, where LFA sensitivity ranged from 28% to 69% ^32-34^. Similarly, a large study of 2,215 people attending a diagnostic centre showed the sensitivity of the Roche and Abbott LFA tests to be only 60.4% and 56.8%, respectively, in detecting RT-PCR-positive (Ct<30) individuals ^35^, which could still be infectious as outlined above. Real-world experience has established that a significant proportion of international air travellers who test positive for SARS-CoV-2 upon arrival are asymptomatic ^36,37^ and would therefore not be reliably detected by lateral flow tests.

Therefore, surveillance testing strategies based solely on such insufficiently sensitive LFA tests may be counterproductive by providing a false sense of security. This was highlighted in an outbreak in Liverpool UK, where 60% of SARS-CoV-2 infections (33% of which had high viral loads) were not detected using LFA. ^38^ A study from the University of Birmingham in April 2021 found that only 5% of SARS-CoV-2-infected individuals were detected by LFA with potentially infectious (Ct<33) viral loads ^39^. In comparison, the test we are reporting here has 98.7% of sensitivity, 97.6% of specificity, and 98% of efficiency at this range of viral loads. The recent experience of an outbreak at the Tokyo Olympics, traced to an individual who had tested negative in an LFA test ^40^, has further highlighted the risks of using LFA tests for asymptomatic surveillance of vulnerable groups.

Rapid SARS-CoV-2 nucleic acid test (NAT) technology, such as the Abbott ID diagnostic test, has therefore been advocated as an alternative or adjunct to LFA tests for COVID-19 surveillance applications, particularly in the next phase of the pandemic ^41^. However, a systematic review of pooled data from the multiple clinical evaluations of the Abbott ID Now test found that its sensitivity was inferior to comparator RT-LAMP tests on crude samples ^42^.

We have prototyped an RT-LAMP-based screening system, that combines sufficient sensitivity for effective viral surveillance with feasible scalability to very high throughput. Given that the viral load in samples from infected but presymptomatic or asymptomatic people can take several days to reach the limit of detection of even the most sensitive tests (such as RT-PCR and RT-LAMP), frequent testing is required to maintain effective viral surveillance ^43^. This is especially important given evidence of SARS-CoV-2 transmission from asymptomatic people with low viral loads, which can gradually increase over several days ^44^, even in vaccinated people ^12^, suggesting that optimal surveillance testing should therefore involve sequential NAT tests.

A recent report of screening 86,000 such samples, with an analytical sensitivity of 87.61% and a specificity of 100% of RT-LAMP in detecting potentially infectious viral loads (Ct < 33) from directly screening saliva clearly showed the value of RT-LAMP for infection surveillance of asymptomatic individuals, if it could be conducted routinely ^45^.

Therefore, we chose to employ an automated RT-LAMP-based screening system in combination with the optimised upstream saliva processing protocol. This way we can provide sufficient sensitivity for effective viral surveillance with feasible scalability to ultra-high throughput screening. Saliva sampling, we propose here, makes repeated testing quicker without compromising the testing quality. Using saliva as a test substance is fundamental for the efficient large scale testing strategy and makes frequent testing feasible.

Given that the viral load in samples from infected but presymptomatic or asymptomatic people can take several days to reach the limit of detection of even the most sensitive tests (such as RT-PCR and RT-LAMP), frequent testing is required to maintain effective viral surveillance ^43^. This is especially important given evidence of SARS-CoV-2 transmission from asymptomatic people with low viral loads, which can gradually increase over several days ^44^, even in vaccinated people ^12^, suggesting that optimal surveillance testing should therefore involve sequential NAT tests.

For example, weekly surveillance testing using saliva-based RT-LAMP has recently been advocated as a feasible means of complementing vaccines to contain the spread of highly infectious pandemic agents such as SARS-CoV-2 ^46^. Travel biosecurity could therefore be enhanced by sequential testing of the passengers both before departure and upon arrival using rapid NAT tests such as RT-LAMP. Using saliva samples instead of nasopharyngeal collection, makes this approach feasible and reliable.

The integrated high-throughput surveillance system described above allows rapid turnaround from convenient saliva sampling combined with highly efficient analysis, which may help facilitate community acceptance of repeated testing. Importantly, the confidentiality of data from tested individuals is also maintained within the Sentinel sample tracking system, based on anonymous digital tokens, ensuring that the instrument’s reporting of test results remains deidentified. Test results reported from the instrument can only be associated with participant identities externally by authorised entities, such as participants themselves or accredited pathology services, who have exclusive access to the required information.

Given limited opportunity to test our system in the field locally (since due to strict border controls, Western Australia experienced no cases of community transmission of SARS-CoV-2 between April 2020 and October 2021), we collaborated with international COVID screening laboratories for the purposes of this study. Extended large-scale deployments of this system will allow further optimization of testing procedures, adapted to specific feedback from different testing environments. Differences in the operations of seaports, airports and sporting venues or ships will therefore likely necessitate refinement to maximise the process efficiency and acceptability. For example, we are now collaborating with resources companies requiring high throughput rapid biosecurity screening measures, to protect their remotely deployed workforce from risks to occupational health and safety imposed by severe infection.

## Summary

The Sentinel surveillance system combines the required components of a sensitive molecular screening system to support efficient detection of all potentially infectious asymptomatic carriers at pandemic-scale. Within the same platform we combined highly parallel sample processing and sensitive biochemistry with the development of random access, continuous flow robotic system.

We have established proof-of-concept for integration of a saliva-based RT-LAMP assay with a surveillance instrument, currently capable of screening up to 4000 reactions per hour with 98% of efficiency. This versatile pathogen vigilance platform offers a unique combination of accuracy and scalability to provide a feasible way to mitigate the risk of viral transmission as borders open and new viral threats arise in the future.

## Methods

### Sample preparation

The samples used in this study were collected from the Sir Charles Gairdner and Osborne Park Health Care Group. All methods were carried out in accordance with relevant guidelines and regulations. The study protocol was approved and endorsed by the Sir Charles Gairdner and Osborne Park Health Care Group Human Research Ethics Committee. Saliva samples were collected, with informed consent, from returned travellers in hotel-quarantine who had been diagnosed as SARS-CoV-2 positive from an approved RT-qPCR diagnostic test (using nasopharyngeal swabs) by the Western Australian state pathology service laboratory (PathWest). None of the individuals required hospitalization. Samples were diluted inan equal volume of viral transport medium (VTM) and stored at - 20 °C. Saliva/VTM was then diluted in AviSal Sample Collection Buffer (Hayat Genetics) and heat inactivated at 95 °C for 10 minutes before being added to Direct RT-qPCR and RT-LAMP reactions.

### Reverse-transcription Loop-mediated isothermal amplification (RT-LAMP)

RT-LAMP primer sequences used in this study have been published elsewhere and are as follows: Zhang E1/N2 (16); Huang O117 and Huang S17 (both Huang primer sets described inin the main text (25)). For internal control reactions, we used rActin primers from NEB.

Oligonucleotides were synthesized by Integrated DNA Technologies, Inc. (IDT, Coralville, IA, USA). F3, B3, LoopF and LoopB primers were desalted after synthesis; FIP and BIP primerswere HPLC-purified. 10x RT-LAMP working primer mixes were prepared in nuclease-free water and contained 2 μM F3, 2 μM B3, 16 μM FIP, 16 μM BIP, 4 μM LoopF and 4 μM LoopB primers. Lyophilized Novacyt S primers (binding to the S-gene in an unknown location, Novacyt Primerdesign, United Kingdom) were reconstituted with a proprietary reconstitution buffer.

Fluorometric RT-LAMP reactions were set up in 96-well PCR plates with WarmStart LAMP 2X Master Mix (E1700, NEB) according to manufacturer’s instructions. Dual-readout (fluorometric and colorimetric) RT-LAMP reactions were set up in 96-well PCR plates with either WarmStart Colorimetric LAMP 2X Master Mix (M1800, NEB) or WarmStart Colorimetric LAMP 2X Master Mix with UDG (M1804, NEB), both supplemented with 1x LAMP Fluorescent Dye (NEB), or Rapid Colorimetric & Fluorometric One Step LAMP SARS-CoV-2 Test Kit (Hayat Genetik, Istanbul, Turkey), according to manufacturer’s instructions. All reactions were overlaid with 15 μl mineral oil (M5904, Sigma-Aldrich, USA). Plates were sealed with clear adhesive films (Microseal B, Bio-Rad Laboratories, CA, USA) and placed at 65 °C on a Sentinel (Avicena Systems, Perth, Australia) in fluorometric mode. The Sentinel was fitted with a 470 nm excitation light source, a 20 Megapixel CMOS camera, and a 500 nm long pass emission filter. Fluorescence detection was performed at 30 sec intervals. Fluorescence signals were converted into arbitrary units by ImageJ software analysis and normalized to the baseline fluorescence signal (set at 7 min after reaction start). Samples were deemed positive for SARS-CoV-2 when fluorescence signal intensity reached a threshold, set as 1.5x baseline fluorescence signal intensity – corresponding to more than 3 standard deviations from the mean baseline signal). A time-to-threshold was calculated for each reaction. Colour change was confirmed by photography with a smartphone at the beginning and end (30 mins) of the reaction and blind-coded. Incubation on ice for 1 to 2 minutes improved colour contrast, making scoring easier. A positive result was scored when colour had changed from pink to orange or yellow.

### RNA extraction and Reverse transcription – quantitative polymerase chain reaction (RT-qPCR)

For RT-qPCR using the PerkinElmer New Coronavirus Nucleic Acid Detection Kit (PerkinElmer, Waltham, MA, USA), RNA was extracted from saliva/VTM using the Chemagic360 (PerkinElmer, Waltham, MA, USA) according to the manufacturer’s protocol, prior to 38 μl of extracted nucleic acid being added to each RT-PCR reaction.

RT-qPCR was also performed on extracted RNA as an accredited Laboratory-DevelopedTest (LDT) by the Western Australian Government pathology service laboratory (PathWest).

Direct RT-qPCR was performed on heat-inactivated saliva/VTM/AviSal using both the PerkinElmer New Coronavirus Nucleic Acid Detection Kit (PerkinElmer, Waltham, MA, USA)and the Real-Time Fluorescent RT-PCR Kit for Detecting SARS-CoV-2 (BGI Europe, Copenhagen, Denmark).

Except for the LDT assays, all amplifications were performed on a QuantStudio7 Pro Real-Time PCR System (Applied Biosystems, Bedford, MA, USA) according to either PerkinElmer’sor BGI’s specified cycling parameters.

### SARS-CoV-2 DNA and RNA templates for LAMP reactions

Initial optimization reactions using N-primers were carried out with 2019-nCoV_N PlasmidControl DNA (Integrated DNA Technologies) as template. The plasmid contains the complete 2019 SARS-CoV-2 nucleocapsid gene. Aliquots of this SARS-coV-2 plasmid control stock solution (200,000 copies/μl) were stored at -20C; dilutions were prepared in nuclease-free waterand freeze-thawed up to 5 times without any observed reduction in the limit of detection (not shown).

Subsequent positive control experiments were carried out with Twist Synthetic SARS-CoV-2 RNA Control 1 (#102019, Australian variant, Lineage B, Twist Bioscience, San Francisco, CA, USA) was used as a positive control in all RT-LAMP assays, while Control RNA 23 (Twist #104533, Delta variant, lineage B1.617.2) was used to confirm detection by RT-LAMP of the “Delta” variant. Control RNA 48 (Twist #105204, Omicron variant, lineage B1.1.529) was used to confirm detection by RT-LAMP of the “Omicron” variant. In all cases, Twist SARS_CoV-2 control RNA consists of six non-overlapping 5000 nucleotide fragments generated by transcription of DNA fragments into ssRNA. The fragments span 99.9% of the viral genome. Control RNA stocks were aliquoted and stored at -80 °C; for each experiment, fresh dilutions were prepared in nuclease-free water and kept at 4 °C for no more than 3 hours.

### Independent comparator study

The samples used in this study were collected as part of an asymptomatic screening programme. Sample information was collected using an app, which blinded the samples to the lab. A study was performed to determine the specificity and sensitivity of the LAMP test by comparing its results to the approved RT-PCR test. Comparator study used 150 genuine positive samples with wide range of viral load and 250 genuine negative samples with wide range of viral load. All samples were analysed by RT-qPCR using ELITE MGB Kit used on an Elite InGenius system and results were compared head-to-head with Avicena and Hayat Genetics LAMP results. Different cut-off times for detection (Time-To-Threshold, TTT) were trialled to determine the effect on sensitivity, specificity, positive predictive value (PPV), negative predictive value (NPV) and efficiency of the assay. The Avicena/Hayat test was compared to the direct LAMP assay used in the UK developed by Optigene Ltd and to a RAT approved for use within the UK (SureScreen Diagnostics).

## Data Availability

All data produced in the present study are available upon reasonable request to the authors

## Author contributions

Conceptualization: SK, DW, PW

Methodology: TH, RD, PO

Investigation: TH, RD, JDR, PO, JMM, MM, PEH

Project administration: SK, PW

Writing – original draft: SK, PW, RD, TH

Writing – review & editing: SK, DW, PW, TH, PO, RD, JMM

Joint-first (RD and TH) and senior (SK and PW) authors contributed equally to those roles in the study.

## Data and materials availability

All data are available in the main text or the supplementary materials.

## Supplementary Materials

Supplementary Text

Figs. S1 to S4

