## Supplementary material for "Validation of a rapid, saliva-based, and ultra-sensitive SARS-CoV-2 screening system for a pandemic-scale infection surveillance": Supplem

**This PDF file includes:**

Figs. S1 to S4

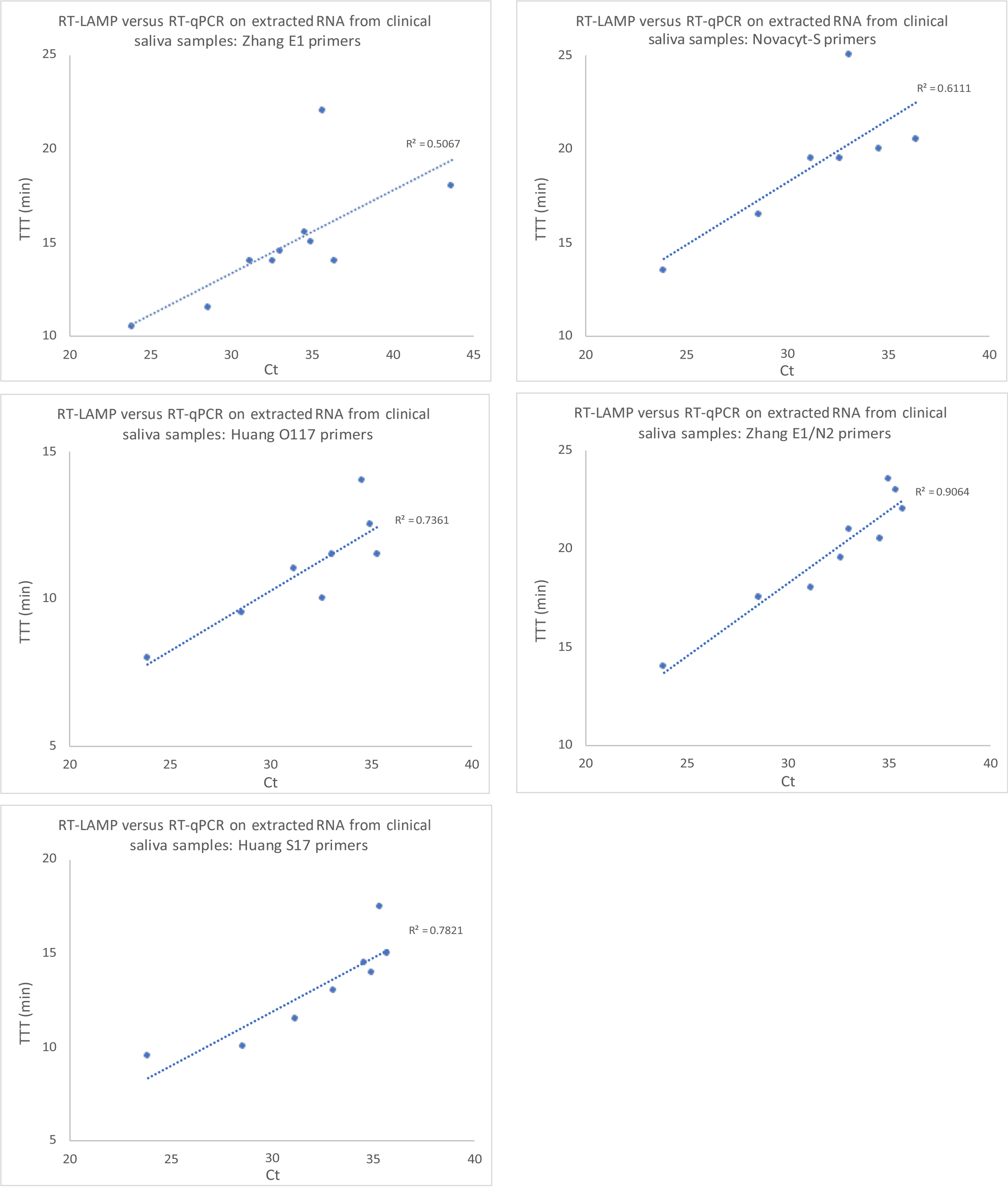


**Suppl. Figure S1: Concordance between RT-LAMP and RT-qPCR**. There was a proportional relationship between Ct values (RT-qPCR) and TTT values for five published RT-LAMP primer sets, demonstrating that RT-LAMP provides a quantitative readout of viral load.

**A**

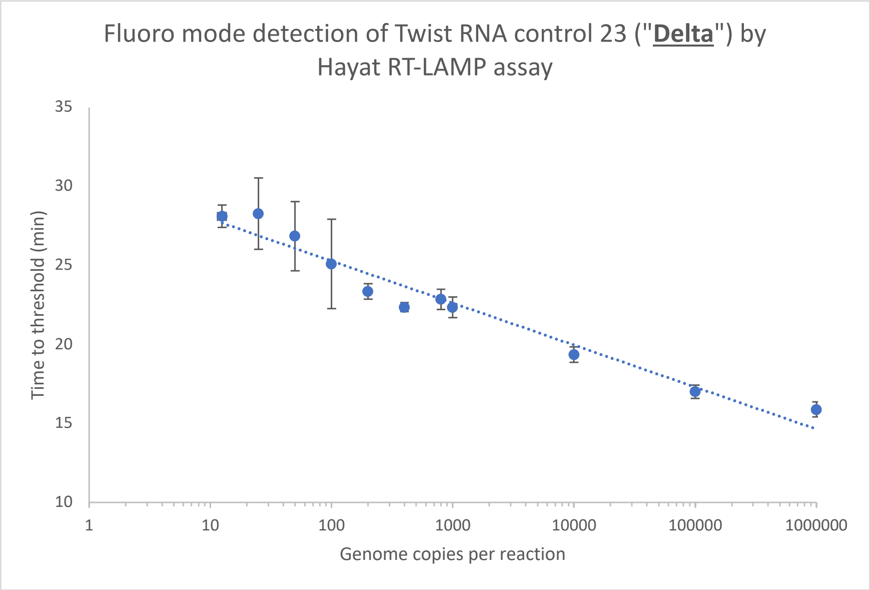


**B**

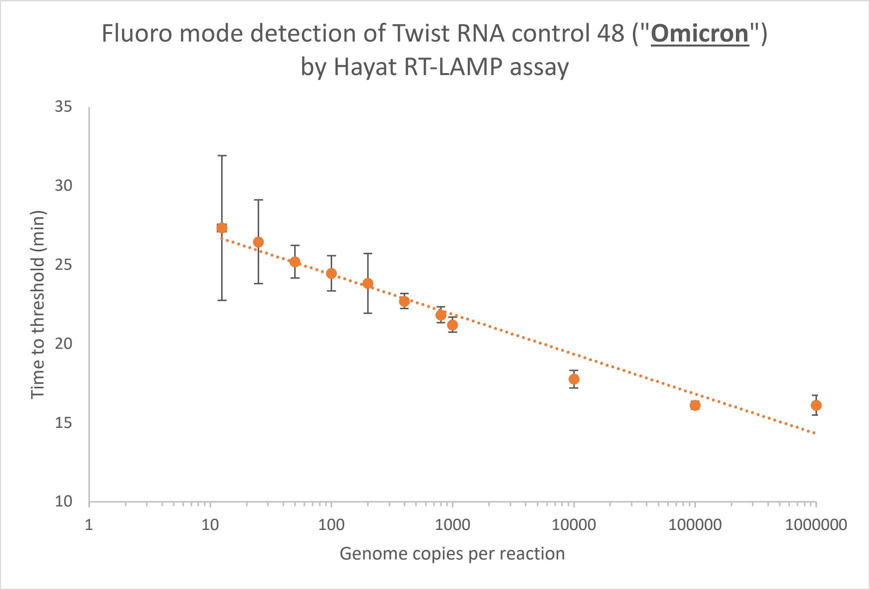


**Suppl. Figure S2. A. Detection of Twist SARS-CoV-2 control RNA 23 ("Delta variant"), and B. Detection of Twist SARS-CoV-2 control RNA 48 ("Omicron variant").** RT- LAMP (Hayat Genetics/Avicena) was performed in a fluorescence mode on a dilution series of Twist Bioscience Control 23 and 48 synthetic RNA. Each datapoint is the mean of four technical replicates. Error bars represent the standard deviation.

| NIBS | VIASURE Ct | Time-to-Threshold [min] | | | | | | | |
|---|---|---|---|---|---|---|---|---|---|
| | | RapiLyze/Optigene | | | | AviSal/Hayat | | | |
| | | rep 1 | rep 2 | rep 3 | Sensitivity | rep 1 | rep 2 | rep 3 | Sensitivity |
| 14 | 18.0 | 16.1 | 12 | 14 | | 18.6 | 16.5 | 21.1 | |
| 4 | 18.2 | 13 | 14 | 18.6 | | 16 | 16 | 17.1 | |
| 10 | 18.2 | 13.5 | 12.5 | 14 | | 16 | 16 | 16 | |
| 26 | 18.2 | 12.5 | 15.1 | 13.5 | | 16 | 16 | 15.5 | |
| 25 | 18.4 | 15.6 | 12.5 | 11 | 100% | 16 | 16.5 | 16 | |
| 5 | 20.7 | 15.1 | 14 | 15.1 | | 17.1 | 16.5 | 16.5 | |
| 18 | 20.8 | 20.1 | 15.6 | 18.6 | | 16.5 | 16.5 | 16.5 | |
| 6 | 20.9 | 14.6 | 13.5 | 16.1 | | 15.5 | 16.5 | 18.6 | |
| 30 | 20.9 | 15.1 | 13.5 | 15.6 | | 16.5 | 17.1 | 16 | |
| 7 | 21.1 | - | 13 | 13 | | 16.5 | 16.5 | 16.5 | 100% |
| 17 | 23.6 | 27.1 | 24.6 | 26.6 | | 18.6 | 22.6 | 20.1 | |
| 19 | 23.7 | - | 25.6 | 19.6 | 78% | 18.6 | 17.6 | 19.6 | |
| 11 | 24.0 | - | 17.6 | 29.1 | | 17.6 | 18.1 | 18.1 | |
| 28 | 24.1 | 20.1 | 18.6 | 20.1 | | 18.6 | 18.1 | 18.6 | |
| 8 | 24.1 | 25.1 | 20.6 | 22.1 | | 18.1 | 21.6 | 18.1 | |
| 9 | 25.2 | - | - | 24.6 | | 19.6 | 19.6 | 19.6 | |
| 12 | 25.2 | - | 29.1 | 25.1 | | 18.1 | 19.1 | 19.6 | |
| 20 | 25.3 | 16.1 | 20.1 | - | | 19.6 | 20.6 | 20.1 | |
| 24 | 25.4 | - | 24.6 | - | | - | - | - | 0% |
| 27 | 25.5 | 29.6 | 19.1 | 19.6 | 44% | 17.1 | 19.1 | 19.1 | |
| 15 | 27.3 | - | 23.1 | - | | 19.6 | 20.6 | 19.1 | |
| 2 | 27.4 | - | 21.1 | 21.6 | | 20.1 | 19.1 | 18.6 | 100% |
| 3 | 28.4 | - | - | 28.6 | | 25.1 | 23.1 | 25.1 | |
| 22 | 29.0 | - | - | 25.1 | | 19.1 | 22.6 | 20.6 | |

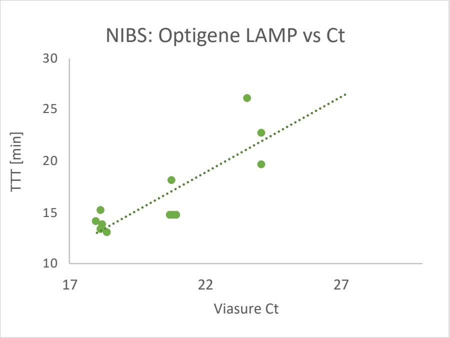


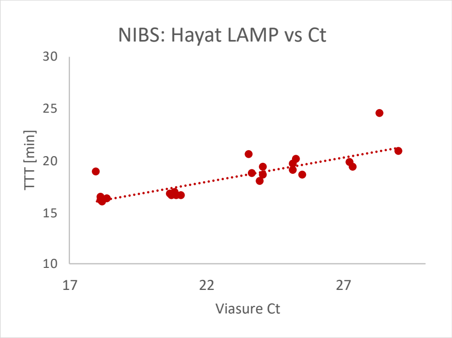


| WHO original concentration | Equivalent concentration | Final concentration | PCR #1 | | Hayat LAMP #1 | PCR #2 | | Hayat LAMP #2 |
|---|---|---|---|---|---|---|---|---|
| IU/ml | Copies/mL | Copies/mL | ORF8 | RdRp | TTP | ORF8 | RdRp | TTP |
| 7.7 | 50,000,000 | 1,500,000 | 22.17 | 21.25 | 12.8 | 21 | 21.96 | 15.5 |
| 6.7 | 5,000,000 | 150,000 | 24.87 | 24.04 | 14.6 | 23.98 | 25.03 | 19.6 |
| 5.7 | 500,000 | 15,000 | 27.9 | 27 | 17.9 | 28.07 | 29.1 | 25.1 |
| 4.7 | 50,000 | 1,500 | 31.59 | 30.79 | 21.4 | 31.15 | 33.79 | 26.1 |
| 3.7 | 5,000 | 150 | 35.07 | 35.25 | - | 35.9 | 35.69 | - |
| 2.7 | 500 | 15 | 41.54 | 37.48 | - | - | - | - |
| 1.7 | 50 | 1.5 | - | - | - | - | - | - |
| 0.7 | 5 | 0.15 | - | - | - | - | - | - |

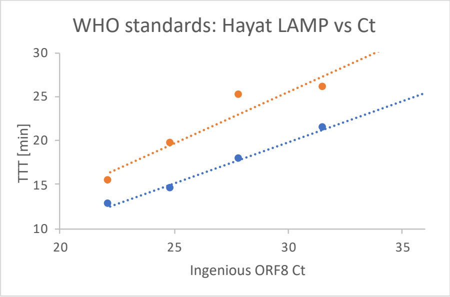


**Suppl. Figure S3. Comparative analysis of NIBSC standardised viral load panel between Hayat and Optigene Direct RT-LAMP assays**

| Sample | Respiratory Pathogen | Hayat LAMP (n=4) |
|---|---|---|
| MRES01 | *Human Metapneumovirus* | *not detected* |
| MRES02 | *Legionella pneumophila* | *not detected* |
| MRES03 | *Adenovirus* | *not detected* |
| MRES04 | *Rhinovirus/ Enterovirus, Picornavirus, Enterovirus* | *not detected* |
| MRES05 | *Parainfluenza 2 & group* | *not detected* |
| MRES06 | *Coronavirus, Coronavirus OC43, RSV, RSV-B* | *not detected* |
| MRES07 | *Bordetella species, Bordetella pertussis, Picornavirus, Rhinovirus, Enterovirus* | *not detected* |
| MRES08 | *Parainfluenza 4 & group* | *not detected* |
| MRES09 | *Chlamydophila pneumoniae* | *not detected* |
| MRES10 | *Human Metapneumovirus* | *not detected* |
| MRES11 | *No Pathogen Detected* | *not detected* |
| MRES12 | *Parainfluenza 1 & group* | *not detected* |
| MRES13 | *Mycoplasma pneumoniae* | *not detected* |
| MRES14 | *No Pathogen Detected* | *not detected* |
| MRES15 | *Parainfluenza 3 & group* | *not detected* |
| MRES16 | *Coronavirus, Coronavirus NL63* | *not detected* |
| MRES17 | *Adenovirus, Influenza A, Influenza A H3* | *not detected* |
| MRES18 | *Picornavirus, Rhinovirus, Rhinovirus/ Enterovirus* | *not detected* |

**Suppl. Figure S4. Detection of the other pathogens by the RT-LAMP test.**
